# An evaluation of global Chikungunya clinical management guidelines – a systematic review

**DOI:** 10.1101/2022.02.23.22271379

**Authors:** Eika Webb, Melina Michelen, Ishmeala Rigby, Andrew Dagens, Dania Dahmash, Vincent Cheng, Reena Joseph, Samuel Lipworth, Eli Harriss, Erhui Cai, Robert Nartowski, Pande Putu Januraga, Keerti Gedela, Evi Sukmaningrum, Helen Groves, Peter Hart, Tom Fletcher, Lucille Blumberg, Peter Horby, Shevin T Jacob, Louise Sigfrid

## Abstract

**Background:** Chikungunya virus (CHIKV) has expanded its geographical reach in recent decades and is an emerging global health threat. CHIKV can cause significant morbidity and lead to chronic, debilitating arthritis in up to 40% of infected individuals, impacting on livelihoods. Prevention, early identification, and clinical management are key for improving outcomes. This review aims to evaluate the availability of inclusive, evidence-based clinical management guidelines for CHIKV in a global context.

**Methods:** Six databases were searched systematically from inception to 14^th^ October 2021 and complemented with a grey literature search until 16^th^ September 2021. We included CMGs providing supportive care and treatment recommendations. Two reviewers independently screened records, extracted data and assessed quality using the AGREE II tool. Findings are presented in a narrative synthesis.

**Results:** Twenty-eight CMGs were included; most were of low-quality (median score 2 out of 7 (range 1-7)). None were produced specifically in a low-income country and 54% (15/28) were produced more than five years ago. There were variations in the CMGs’ guidance on the management of different at-risk populations, long-term sequelae, and the prevention of disease transmission in community and hospital settings. In the acute phase, 54% (15/28) recommended hospitalisation for severe cases, however only 39% (11/28) provided clinical management guidance for severe disease. Further, 46% (13/28) advocated for steroids in the chronic phase, yet 18% (5/28) advised against its use.

**Conclusion:** There was a lack of high-quality CMGs that provided supportive care and treatment guidance; this scarcity may impact patient care and outcomes. It is essential that existing guidelines are updated and adapted to provide detailed evidence-based treatment guidelines for different at-risk populations. This study also highlights a need for more research into the management of the acute and chronic phases of CHIKV infection to inform evidence-based care.

Systematic review registration: PROSPERO CRD42020167361

**What was known before:**

- CHIKV is endemic across most of the southern hemisphere, with risk of expansion into new regions driven by global travel, trade, and climate change.
- Infection can result in severe illness with long-term sequelae, particularly in vulnerable groups. Chronic sequelae of CHIKV infection is a cause of significant debilitating morbidity affecting individual functionality and quality of life with wider health system and socio-economic impact.
- There is no effective vaccine or targeted treatment against Chikungunya and supportive care is the mainstay of treatment.
- Even with a limited evidence base, clinical management guidelines (CMGs) are key tools for standardising best available evidence-based care, and reduce inappropriate use of treatments, to reduce morbidity and improve patient outcomes.

**What this study adds:**

- This review highlights a global scarcity of CMGs for chikungunya providing detailed guidance on optimal supportive care and treatment for different at-risk populations and settings.
- There was limited guidance available on care for severe cases, and available guidance was heterogenous and discordant (e.g., on use of analgesia, corticosteroids, and monitoring).
- The limited availability of up-to-date CMGs and heterogenous recommendations identified is a concern, which may impact on equity in access to best available evidence-based care and patient outcomes.
- Further research into access to and implementation of CMGs in different settings is needed, to ensure equitable access to best available care.
- This study also highlights a need for further investment into research into supportive care and treatment for different at-risk populations, and new evidence incorporated into guidelines to reduce morbidity and improve long term outcomes for the people affected by and at risk of Chikungunya.

## INTRODUCTION

Chikungunya is a disease caused by the chikungunya virus (CHIKV); an arthropod-borne virus transmitted to humans primarily by *Aedes* mosquitoes. Since its description in 1952, CHIKV has caused millions of human infections in Africa, the Indian Ocean islands, Asia, Europe, and the Americas. ^1^ A major outbreak in 2004 affected more than 100 countries with over 10 million cases and this was followed by another large outbreak in 2013 in the Americas. ^2^ Multiple factors have contributed to these outbreaks including a lack of vaccines and effective treatments, limited mosquito control in densely populated urban areas, and climate changes.^3^ It is estimated that 1.3 billion people live in areas at risk of CHIKV infection.^4, 5^ Modelling studies of climate change, indicate that regions such as Europe are at risk of introduction of CHIKV in the future.^4, 6, 7^ The recent and projected expansion in geographical range and risk of travel-imported infections, with risk of local outbreaks during favourable transmission conditions have increased CHIKV’s recognition as an emerging global health threat.

Chikungunya has a wide spectrum of clinical presentations, which can be classified into three phases (acute, sub-acute and chronic).^8^ Acute CHIKV (classed as less than three weeks post onset of symptoms) manifests as a febrile illness with predominant symptoms of polyarthralgia, rash, and headache. This can be followed by a subacute phase lasting up to three months.^9^ An estimated 40% of people are affected by long-term chronic sequelae defined as persistent symptoms more than 3 months post-onset, for up to 6 years.^9, 10^ The chronic manifestations of CHIKV include debilitating symptoms of chronic arthralgia, arthritis, fatigue which may lead to disability and diminished quality of life,^11^ with severe impact on an individual’s ability to work. Although the acute infection is self-limiting and rarely life-threatening, it can result in severe illness and mortality particularly in neonates, older adults (over 65 years) and people with comorbidities. ^2, 8, 12–14^ To date, there is no specific treatment approved for acute or chronic chikungunya illness; although vaccines have been developed and tested in humans, none are yet available.^15, 16^ Thus, to improve patient outcomes, and reduce the chronic burden; supportive care is essential for the clinical management of chikungunya.^17^

Parallels can be drawn between CHIKV, and SARS-CoV-2 given that both viruses can cause acute illness followed by long-term sequelae in survivors, which can have a devastating impact on individuals’ psychological and physical health and capacity to return to work. Accordingly, public health interventions adopted by many countries to slow the spread of COVID-19 (i.e., reduction in the number of regular household surveys; diversion of resources towards the COVID-19 response; lockdowns) may likely have had a negative impact on vector surveillance and control.^18, 19^ As we are transitioning out of the pandemic, we need to prepare to shift resources to identify and mitigate the wider pandemic consequences and strengthen our capacity to respond to future epidemics.

CHIKV infection is a serious global public health problem, predominantly affecting populations and health systems in lower resourced settings, and with risk of importation into new, naïve regions. The aim of this review is to explore the availability and accessibility of evidence-based clinical management guidelines for chikungunya globally and evaluate their quality and harmonisation of treatment recommendations.

## METHODS

We conducted a systematic review of chikungunya CMGs using Cochrane systematic review methodologies,^20^ structured according to the Preferred Reporting Items for Systematic Reviews and Meta-Analyses (PRISMA) statement (supplementary file).^21^ The protocol is registered with the International Prospective Register of Systematic Reviews (PROSPERO) (CRD42020167361).^22^

### Search strategy

We conducted an electronic database search for chikungunya CMGs using Ovid Medline, Ovid Embase, Ovid Global Health, Scopus, Web of Science Core Collection and WHO Global Index Medicus from inception to 14^th^ October 2021. Search strategies applied the Canadian Agency for Drugs and Technologies in Health (CADTH) database guidelines search filter (supplementary file 1).^23^ Further CMGs were identified through an extensive systematic grey literature search up to 16^th^ September 2021. We searched Google and Google Scholar using predefined keywords in main languages including Arabic, English, French, German, Mandarin, Russian, and Spanish. We also contacted clinician members of the global International Severe Acute Respiratory and Emerging Infection Consortium (ISARIC) network requesting CMGs.^24^

### Eligibility criteria

We included chikungunya CMGs that provided treatment and/or supportive care recommendations. There was no language limitations We excluded CMGs that were pure diagnostics, animal, or public health guidelines. Only the most recent version of any CMG was included.

### Screening and data extraction

Search results were screened independently by two reviewers using the Rayan systematic review software.^25^ CMGs were screened first by title and abstract, then by full text. Screening, data extraction and critical appraisal were completed by two reviewers. Data was extracted by one reviewer using a standardised form and validated by a second reviewer (supplementary file). We extracted data on bibliography, issuing organisations, populations covered, supportive care and treatment recommendations for the acute and chronic disease (supplementary file). Disagreements were resolved via consensus or by a third reviewer. Non-English language CMGs were screened, and data was translated using Google translate and screened and data extracted by reviewers with good to excellent knowledge of the language.

### Quality appraisal

Quality was independently assessed by two reviewers using the Appraisal of Guidelines for Research and Evaluation II (AGREE II) Instrument.^26^ This tool provides an objective framework which aims to assess the guideline development process and quality. It is s a 23-item tool that spans six domains comprising different aspects of the CMG: 1) scope and purpose; 2) stakeholder involvement; 3) rigour of development; 4) clarity of presentation; 5) applicability and 6) editorial independence. Each domain has several sub-criteria which are scored to assess whether the criteria are met using a seven-point Likert scale, from 1 (strongly disagree) to 7 (strongly agree). A score of 100% is achieved if a CMG scores 7 for all items in the domain and 0% if each reviewer scored 1 for all domain items. The final domain score was calculated as the average score by the different reviewers, as per the AGREE-II tool’s user manual.^26^ When there was limited information about the methodology presented in the CMG, efforts were made to search for additional information via associated webpages.

CMGs were considered of high quality if they scored more than 60% in three or more domains, including domain three (rigour of development), which is considered a high-quality indicator; moderate quality if they scored more than 60% in three or more domains but not in domain three and low quality if they did not reach these criteria. As per AGREE-II methodology, individual CMGs were also given an overall quality assessment score, which was informed by the domain scores, ranging from one to seven (high-quality score ≥6; medium-quality score 4-5; low-quality score ≤ 3), together with a recommendation for use with or without further modifications.^26^ CMGs with a total overall quality score of 1 were not recommended for use, total overall scores of 2-5 were recommended for use with modifications and 6-7 recommended for use without modifications.

### Data analysis

We conducted a narrative synthesis of the quality, availability, scope, and inclusivity of the CMGs. The availability of CMGs was assessed by whether open-sourced CMGs could be identified and were stratified by origin: (1) international and regional organisations (e.g., WHO; PAHO); (2) national organisations (e.g., Ministries of Health) and (3) clinical reference websites (e.g., Medscape, UptoDate). We assessed inclusivity based on inclusion of recommendations for traditionally more vulnerable at-risk groups, including infants/children, pregnant women, older people, and people living with HIV or comorbidities that may render them at higher risk of severe disease. The ggplot2 library and Tableau software were used to produce graphics. ^27, 28^

### Patient public involvement

There was no patient or public involvement due to the ongoing pandemic restrictions

## RESULTS

From 2981 records screened, twenty-eight CMGs met the inclusion criteria (Figure 1). ^9, 29–54^

**Figure 1:**
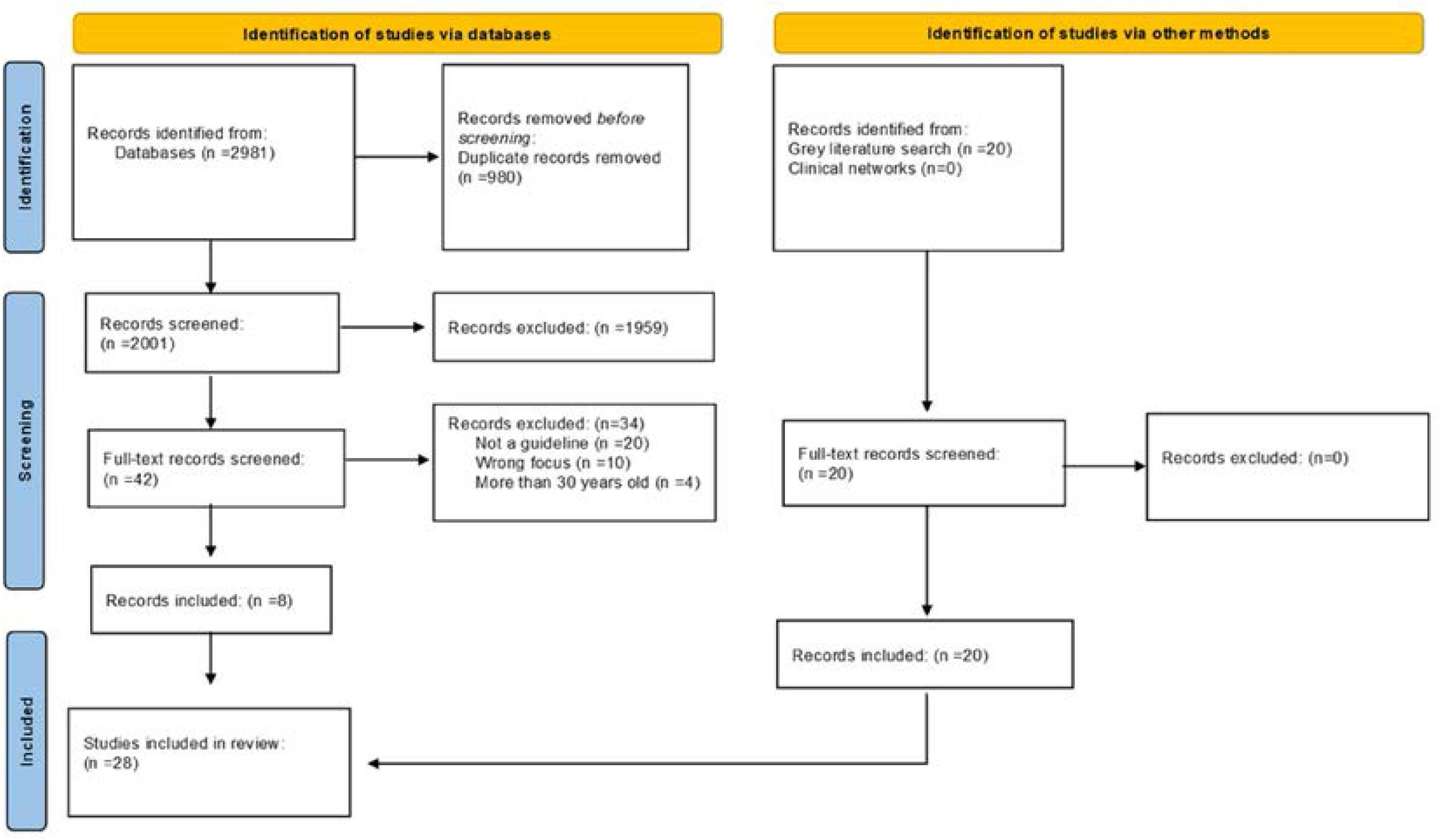
PRISMA diagram.

### Characteristics of included CMGs

Half of the CMGs were in English (50%, 14/28); ^9, 30–32, 34, 36, 37, 42, 44–47, 52, 54^ 43% (12/28) in Spanish ^29, 35, 38– 41, 43, 48–50, 53, 55^ and 7% (2/28) in Portuguese.^33, 51^ Most were produced by National Health Organisations (61%, 17/28), and 46% (13/28)^32–34, 36, 37, 39, 44–46, 51–54^ published in the last 5 years (Table 1).

**Table 1.** AGREE II scores. The table present the results of the assessment of each Chikungunya clinical management guidelines using the AGREE II scores by domain and the overall quality.

| CMG | Year | Scope and purpose (%) | Stakeholder involvement (%) | Rigour of development (%) | Clarity of presentation (%) | Applicability (%) | Editorial Independence (%) | Overall quality (1-7) |
| --- | --- | --- | --- | --- | --- | --- | --- | --- |
| ACCAR | 2018 | 61.1 | 36.1 | 44.8 | 55.6 | 20.8 | 41.7 | 1 |
| BCDC | 2017 | 19.4 | 11.1 | 13.5 | 61.1 | 12.5 | 16.7 | 2 |
| BMS | 2015 | 47.2 | 27.8 | 18.8 | 72.2 | 27.1 | 16.7 | 1 |
| BSR | 2017 | 44.4 | 22.2 | 51 | 69.4 | 10.4 | 45.8 | 2 |
| BZLMS | 2017 | 55.6 | 30.6 | 28.1 | 58.3 | 45.8 | 0 | 1 |
| CDC | 2020 | 8.3 | 25 | 7.3 | 58.3 | 12.5 | 0 | 1 |
| CMS | 2018 | 69.4 | 19.4 | 24 | 69.4 | 39.6 | 0 | 2 |
| CRMS | 2014 | 47.2 | 30.6 | 8.3 | 50 | 50 | 0 | 1 |
| DRMSP | 2014 | 83.3 | 41.7 | 35.4 | 61.1 | 62.5 | 0 | 4 |
| EMS | 2014 | 58.3 | 30.6 | 20.8 | 66.7 | 52.1 | 0 | 2 |
| ESMS | 2014 | 77.8 | 38.9 | 26 | 69.4 | 29.2 | 12.5 | 3 |
| GMS | 2015 | 69.4 | 50 | 16.7 | 58.3 | 16.7 | 0 | 2 |
| IMOH | 2016 | 16.7 | 5.6 | 9.4 | 58.3 | 12.5 | 16.7 | 1 |
| JIMA | 2020 | 52.8 | 38.9 | 40.6 | 58.3 | 8.3 | 95.8 | 2 |
| MMS | 2015 | 94.4 | 63.9 | 93.8 | 91.7 | 62.5 | 87.5 | 7 |
| MS | 2019 | 8.3 | 44.4 | 36.5 | 58.3 | 4.2 | 41.7 | 1 |
| PAHO | 2011 | 52.8 | 47.2 | 22.9 | 52.8 | 31.3 | 25 | 1 |
| PHE | 2014 | 2.8 | 2.8 | 0 | 27.8 | 4.2 | 0 | 1 |
| PMS | 2015 | 80.6 | 44.4 | 36.5 | 83.3 | 29.2 | 0 | 3 |
| PMSP | 2015 | 75 | 44.4 | 27.1 | 47.2 | 41.7 | 0 | 2 |
| PRMS | 2014 | 27.8 | 13.9 | 27.1 | 72.2 | 31.3 | 0 | 2 |
| PUK | 2014 | 38.9 | 36.1 | 46.9 | 44.4 | 6.3 | 66.7 | 2 |
| RSMBT | 2020 | 55.6 | 33.3 | 22.9 | 24 | 72.2 | 22.9 | 2 |
| SMOH | 2016 | 72.2 | 38.9 | 31.3 | 63.9 | 29.2 | 0 | 3 |
| SPILF | 2015 | 55.6 | 38.9 | 45.8 | 61.1 | 16.7 | 33.3 | 2 |
| UTD | 2020 | 47.2 | 44.4 | 85.4 | 80.6 | 35.4 | 91.7 | 6 |
| WHO | 2017 | 69.4 | 33.3 | 43.8 | 55.6 | 37.5 | 25 | 2 |
| WHOSEA | 2008 | 80.6 | 61.1 | 52.1 | 88.9 | 22.9 | 0 | 4 |
| **Median** | - | **55.6** | **36.1** | **27.6** | **61.1** | **28.1** | **14.6** | **2** |
| **Range** | - | **(8-94)** | **(2-63)** | **(0-93)** | **(27-91)** | **(4-62)** | **(0-87)** | **(0-7)** |

**Abbreviations:** ACCAR: Pan-American League of Associations for Rheumatology-Central American Caribbean and Andean Rheumatology Association, *BCDC*: Bangladesh Centre for Disease Control, *BMS*: Bolivia Ministerio De Salud, *BSR*: Brazilian Society of Rheumatology, *BZLMS*: Brasil Ministério da Saúde, *CDC*: Centers for Disease Control and Prevention, *CMS*: Chile Ministerio De Salud, *CRMS*: Costa Rica Ministerio De Salud, *DRMSP*: República Dominicana Ministerio de Salud Pública, *EMS*: Ecuador Ministerio De Salud, *ESMS*: El Salvador Ministerio De Salud, *GMS*: Guatemala Ministerio De Salud, *IMOH*: India Ministry of Health, *JIMA*: Journal of Indian Medical Association. *MMS*: Mexico Ministerio De Salud, *MS*: Medscape, *PAHO*: Pan American Health Organisation, *PHE*: Public Health England, *PMS*: Peru Ministerio De Salud, *PMSP*: Paraguay Ministerio de Salud, *PRMS*: Puerto Rico Ministerio De Salud, *PUK*: Patient UK, *RSMBT*: Revista da Sociedade Brasileira de Medicina Tropical, *SMOH*: Spain Ministry of Health, *SPILF*: Société de Pathologie Infectieuse de Langue Française, *UTD*: UptoDate, *WHO*: World Health Organisation, *WHOSEA*: World Health Organisation Southeast Asia, *CMG*: Clinical management guidelines, *AGREE*- Appraisal of guidelines for research and evaluation

### Availability

Half of the CMGs were produced in Latin Americas (50%, 14/28),^29, 33, 35, 38, 40, 41, 43, 48–51, 53–55^ 14% in Europe (4/28),^9, 30, 31, 39^ 14% in Asia (4/28),^37, 42, 44, 45^ 11% in North America (3/28) ^32, 34, 36^ and 11% by global organisations (3/28) ^32, 34, 52^ (Figure 2). Most were produced by organisations in high- or upper-middle income countries (61%, 17/28),^33, 35, 38, 40, 41, 48, 49, 51, 54, 55^ followed by lower-middle-income countries (18%, 5/28). ^29, 37, 43–45^ None were produced in a low-income country. (World Bank) (Table 1). Sixty-four percent (18/28) of CMGs were produced in countries where Chikungunya is endemic.

**Figure 2:**
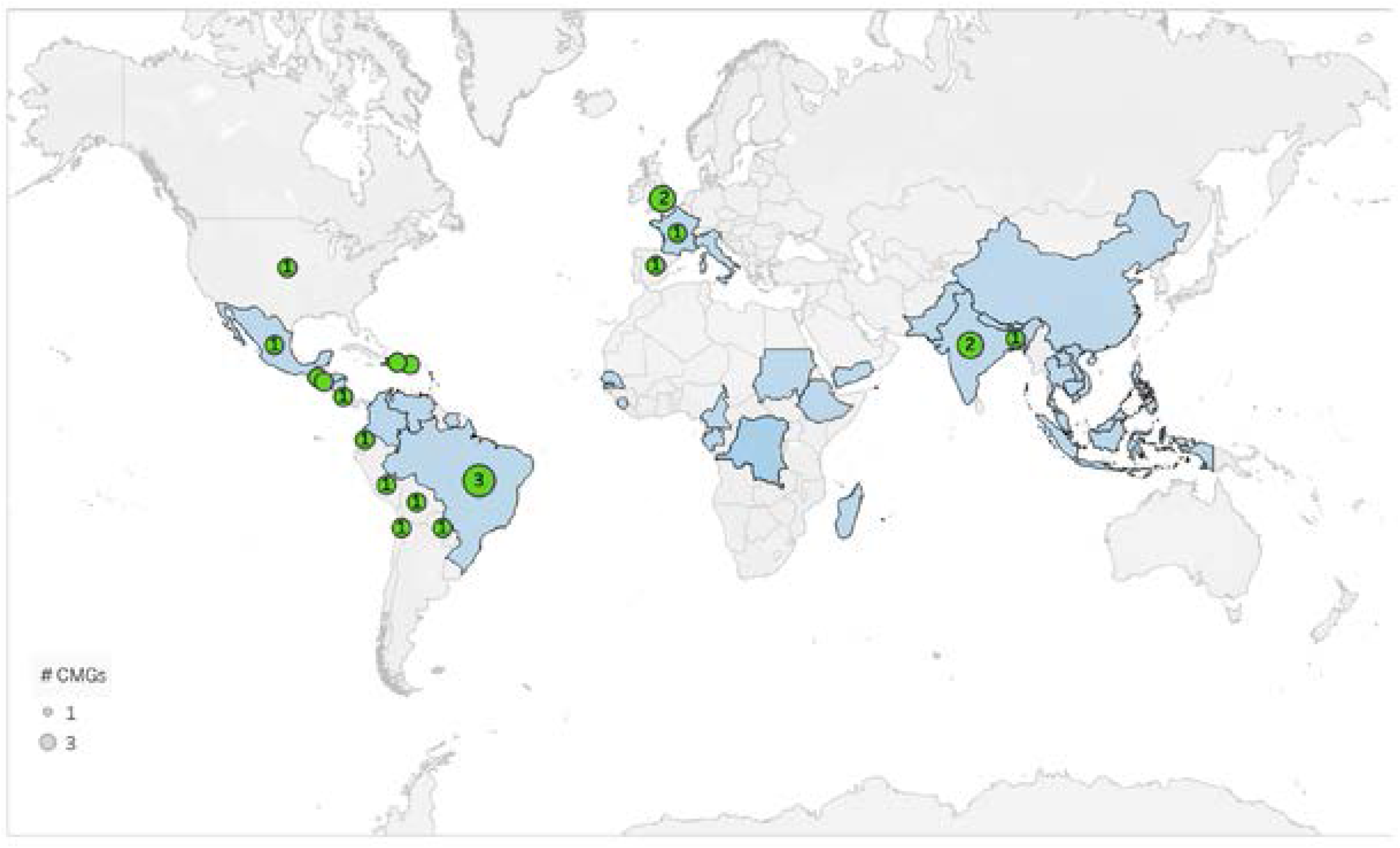
Chikungunya outbreaks (1999-2020) and geographic distribution of identified CMGs.

### Quality

The overall quality of the CMGs ranged from one to seven (median: 2 out of 7) (Table 2). Most (86%, 24/28) were of low quality (score ≤ 3), two of median (scores 4-5) and two of high quality (score 6-7) . The higher scoring CMGs were produced by Mexico Ministerio De Salud, UpToDate, World Health Organisation Southeast Asia and República Dominicana Ministerio de Salud Pública. Although UpToDate produces CMGs for unspecified settings, these are only accessible upon subscriptions. No freely accessible, global, high-quality CMG was identified within our study. The highest scoring domains were clarity of presentation [median (IQR): 61% (58-72)] and scope and purpose [median (IQR): 56% (43-70)]. The lowest scoring domain was editorial independence [median (IQR): 15% (0-35)]. Similarly, the domains for rigour of development [median (IQR): 28% (21-45)], applicability [median (IQR): 29% (16-40)] and stakeholder involvement [median (IQR): 36.1 (2-63)] scored low. Broad variation in scores were seen in the domains for ‘editorial independence’ and ‘rigour of development’ especially (Table 1 and Figure 3).

**Figure 3.**
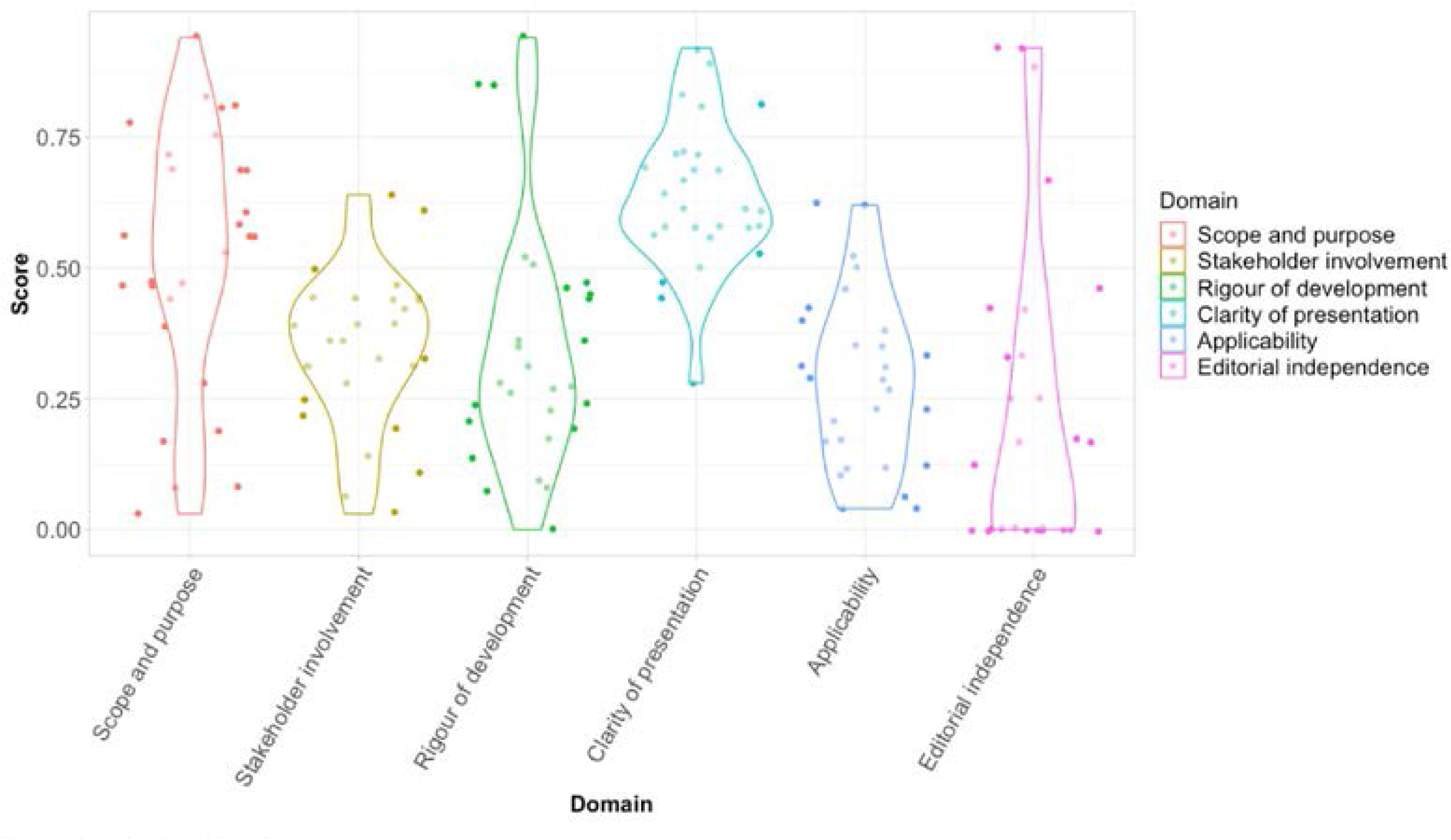
AGREE II domain scores.

**Table 2.**
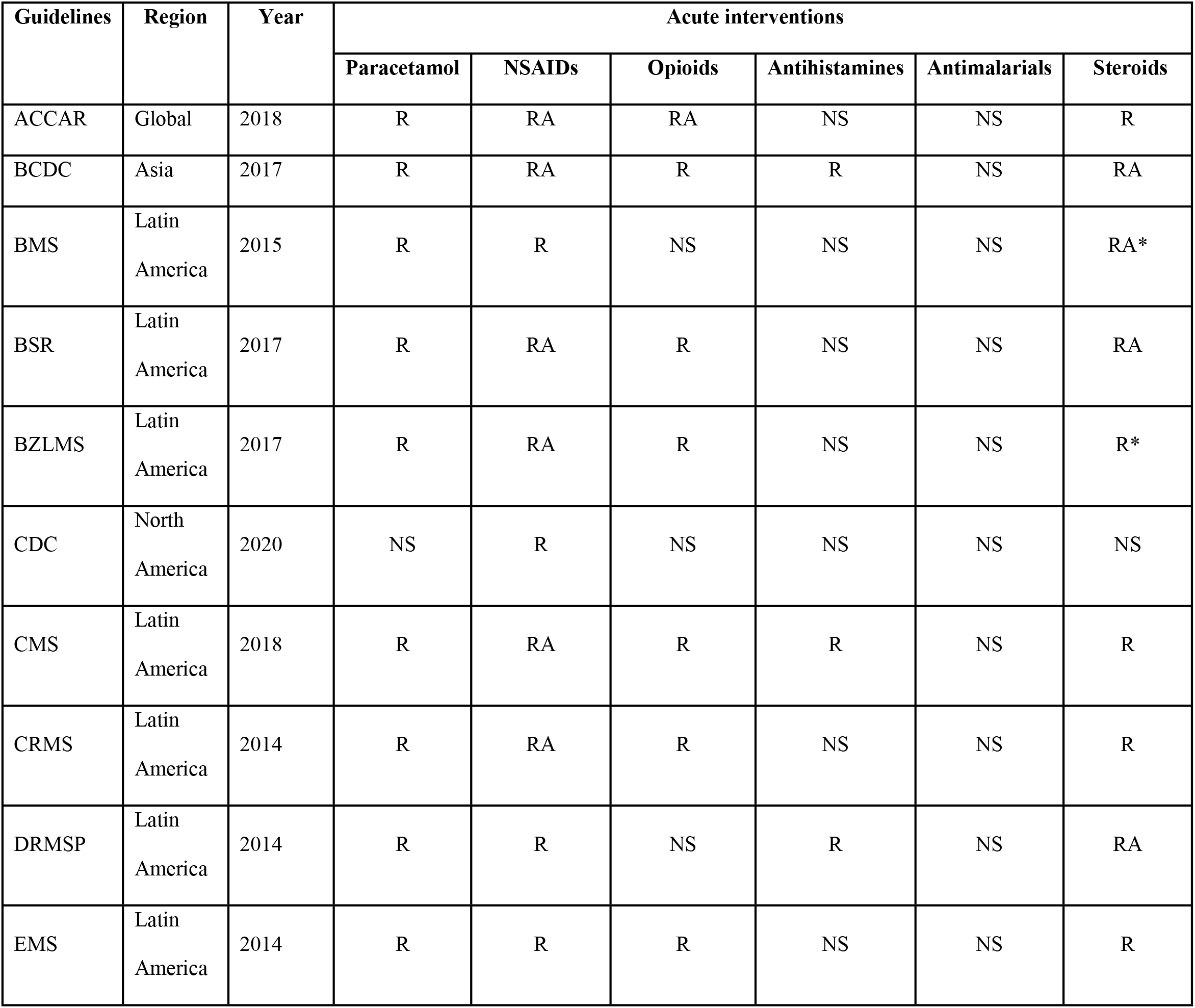
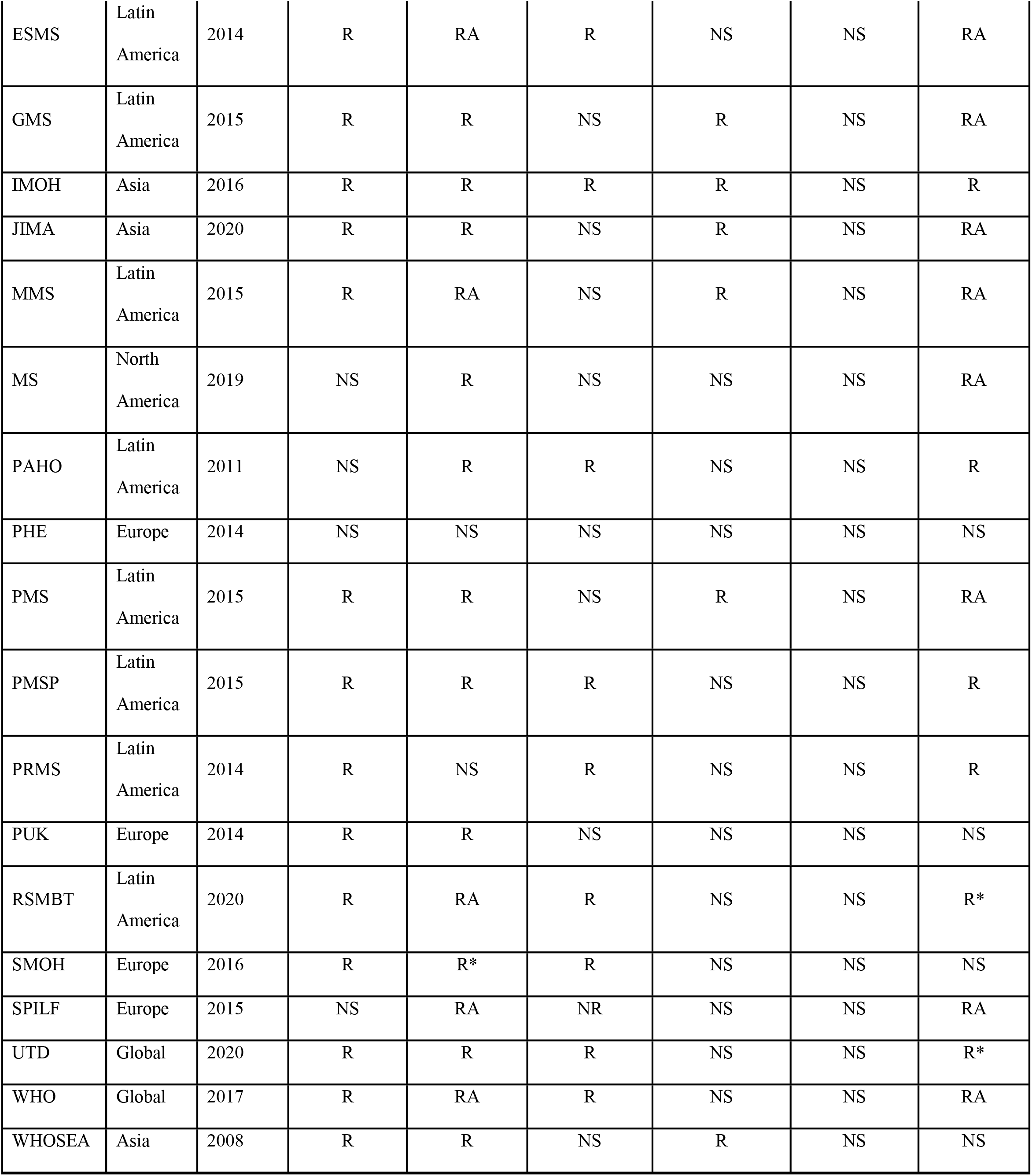
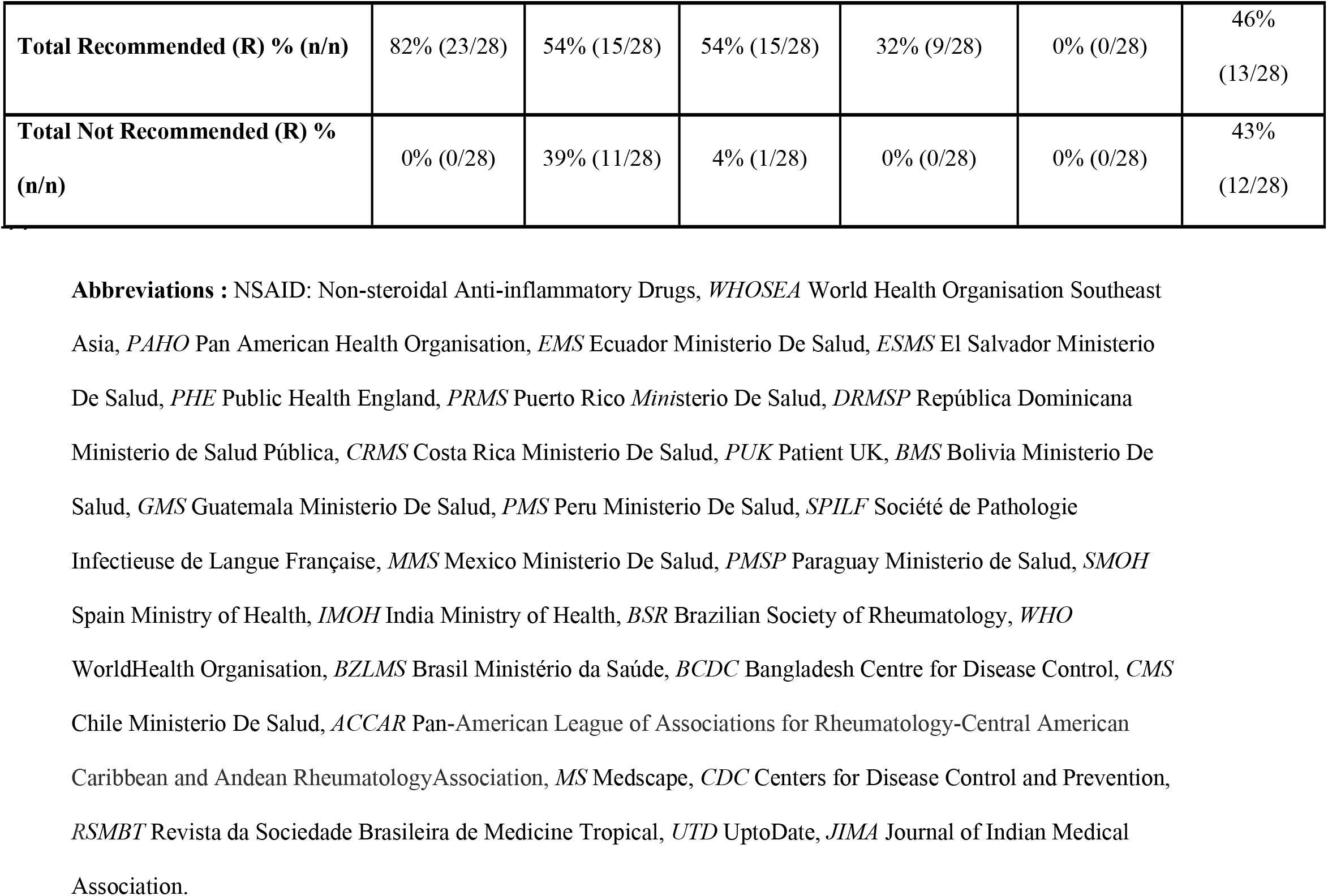
Summary of CMG treatment recommendations in the acute phase. The table presents an overview of the main treatments recommended in the acute phase in each guideline, and if a treatment was recommended to use, not recommended or if no advice was provided. R= recommended. RA= recommended to avoid. NS= not stated. *Indicated in subacute CHIKV

| Guidelines | Region | Year | Acute interventions |  |  |  |  |  |
| --- | --- | --- | --- | --- | --- | --- | --- | --- |
|  |  |  | Paracetamol | NSAIDs | Opioids | Antihistamines | Antimalarials | Steroids |
| ACCAR | Global | 2018 | R | RA | RA | NS | NS | R |
| BCDC | Asia | 2017 | R | RA | R | R | NS | RA |
| BMS | Latin America | 2015 | R | R | NS | NS | NS | RA* |
| BSR | Latin America | 2017 | R | RA | R | NS | NS | RA |
| BZLMS | Latin America | 2017 | R | RA | R | NS | NS | R* |
| CDC | North America | 2020 | NS | R | NS | NS | NS | NS |
| CMS | Latin America | 2018 | R | RA | R | R | NS | R |
| CRMS | Latin America | 2014 | R | RA | R | NS | NS | R |
| DRMSP | Latin America | 2014 | R | R | NS | R | NS | RA |
| EMS | Latin America | 2014 | R | R | R | NS | NS | R |
| ESMS | Latin America | 2014 | R | RA | R | NS | NS | RA |
| GMS | Latin America | 2015 | R | R | NS | R | NS | RA |
| IMOH | Asia | 2016 | R | R | R | R | NS | R |
| JIMA | Asia | 2020 | R | R | NS | R | NS | RA |
| MMS | Latin America | 2015 | R | RA | NS | R | NS | RA |
| MS | North America | 2019 | NS | R | NS | NS | NS | RA |
| PAHO | Latin America | 2011 | NS | R | R | NS | NS | R |
| PHE | Europe | 2014 | NS | NS | NS | NS | NS | NS |
| PMS | Latin America | 2015 | R | R | NS | R | NS | RA |
| PMSP | Latin America | 2015 | R | R | R | NS | NS | R |
| PRMS | Latin America | 2014 | R | NS | R | NS | NS | R |
| PUK | Europe | 2014 | R | R | NS | NS | NS | NS |
| RSMBT | Latin America | 2020 | R | RA | R | NS | NS | R* |
| SMOH | Europe | 2016 | R | R* | R | NS | NS | NS |
| SPILF | Europe | 2015 | NS | RA | NR | NS | NS | RA |
| UTD | Global | 2020 | R | R | R | NS | NS | R* |
| WHO | Global | 2017 | R | RA | R | NS | NS | RA |
| WHOSEA | Asia | 2008 | R | R | NS | R | NS | NS |

|  |  |  |  |  |  |  |
| --- | --- | --- | --- | --- | --- | --- |
| **Total Recommended (R) % (n/n)** | 82% (23/28) | 54% (15/28) | 54% (15/28) | 32% (9/28) | 0% (0/28) | 46%<br>(13/28) |
| **Total Not Recommended (R) %**<br>**(n/n)** | 0% (0/28) | 39% (11/28) | 4% (1/28) | 0% (0/28) | 0% (0/28) | 43%<br>(12/28) |

**Abbreviations :** NSAID: Non-steroidal Anti-inflammatory Drugs, *WHOSEA* World Health Organisation Southeast Asia, *PAHO* Pan American Health Organisation, *EMS* Ecuador Ministerio De Salud, *ESMS* El Salvador Ministerio De Salud, *PHE* Public Health England, *PRMS* Puerto Rico Ministerio De Salud, *DRMSP* República Dominicana Ministerio de Salud Pública, *CRMS* Costa Rica Ministerio De Salud, *PUK* Patient UK, *BMS* Bolivia Ministerio De Salud, *GMS* Guatemala Ministerio De Salud, *PMS* Peru Ministerio De Salud, *SPILF* Société de Pathologie Infectieuse de Langue Française, *MMS* Mexico Ministerio De Salud, *PMSP* Paraguay Ministerio de Salud, *SMOH* Spain Ministry of Health, *IMOH* India Ministry of Health, *BSR* Brazilian Society of Rheumatology, *WHO* WorldHealth Organisation, *BZLMS* Brasil Ministério da Saúde, *BCDC* Bangladesh Centre for Disease Control, *CMS* Chile Ministerio De Salud, *ACCAR* Pan-American League of Associations for Rheumatology-Central American Caribbean and Andean Rheumatology Association, *MS* Medscape, *CDC* Centers for Disease Control and Prevention, *RSMBT* Revista da Sociedade Brasileira de Medicina Tropical, *UTD* UptoDate, *JIMA* Journal of Indian Medical Association.

**Table 3.**
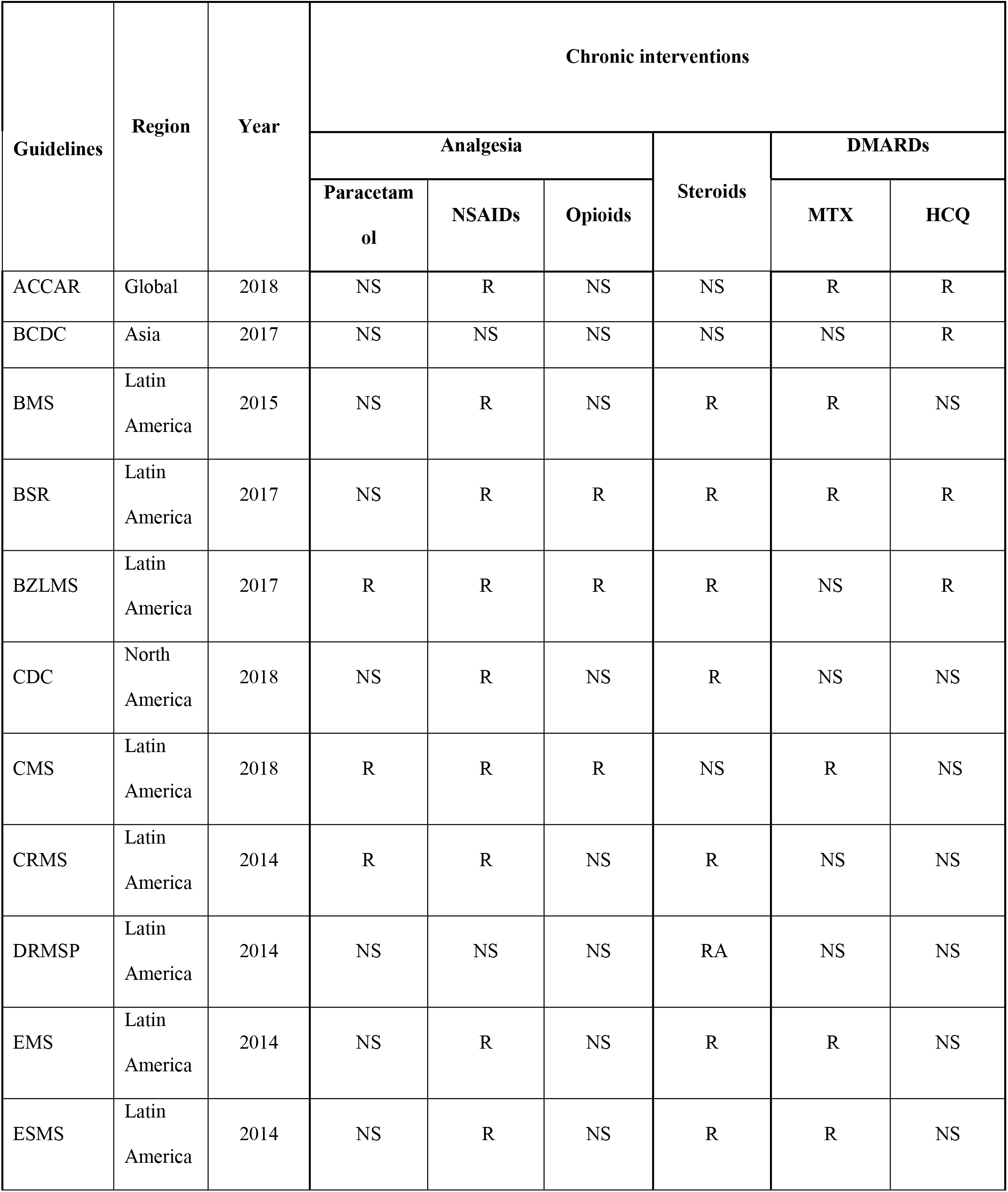
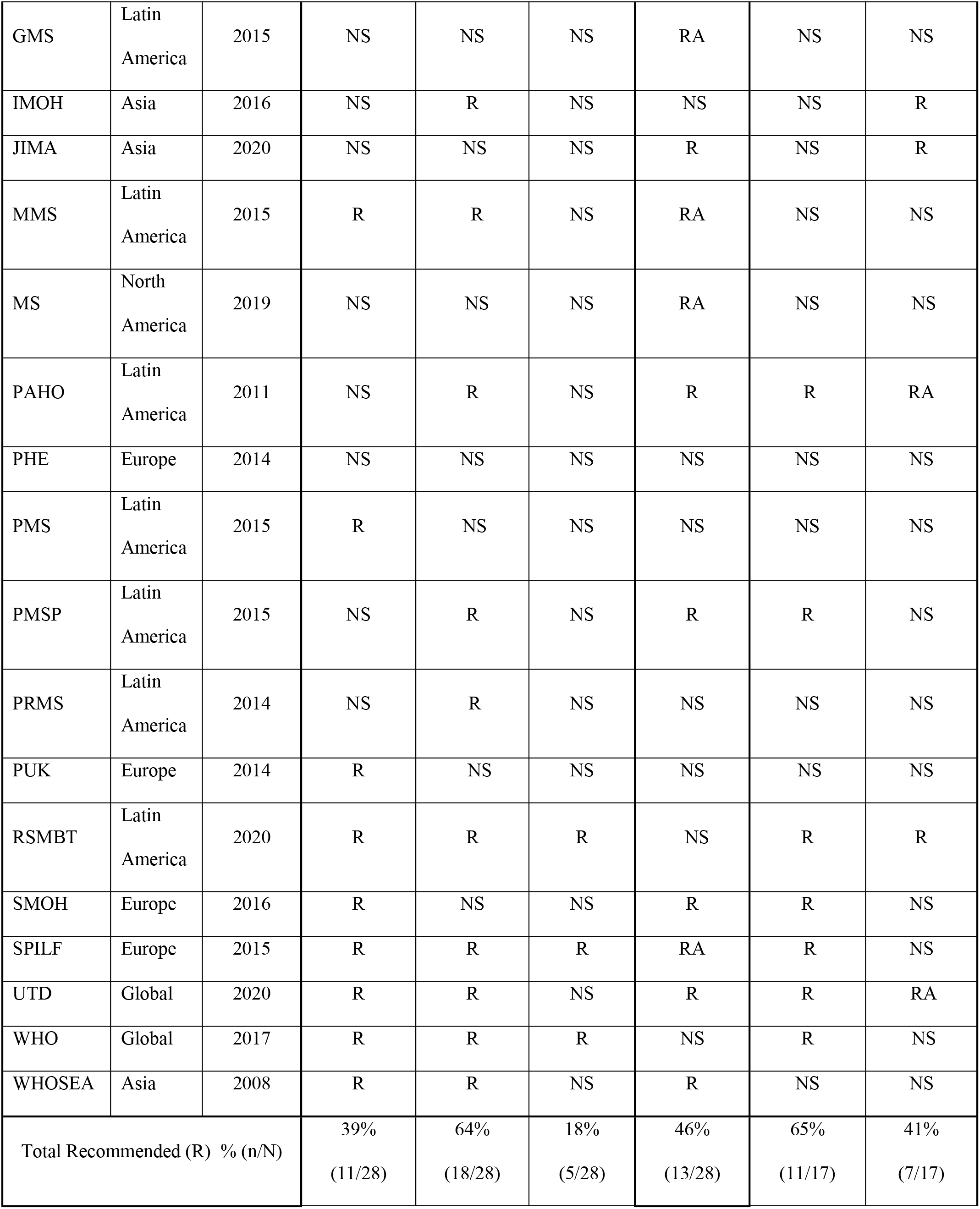
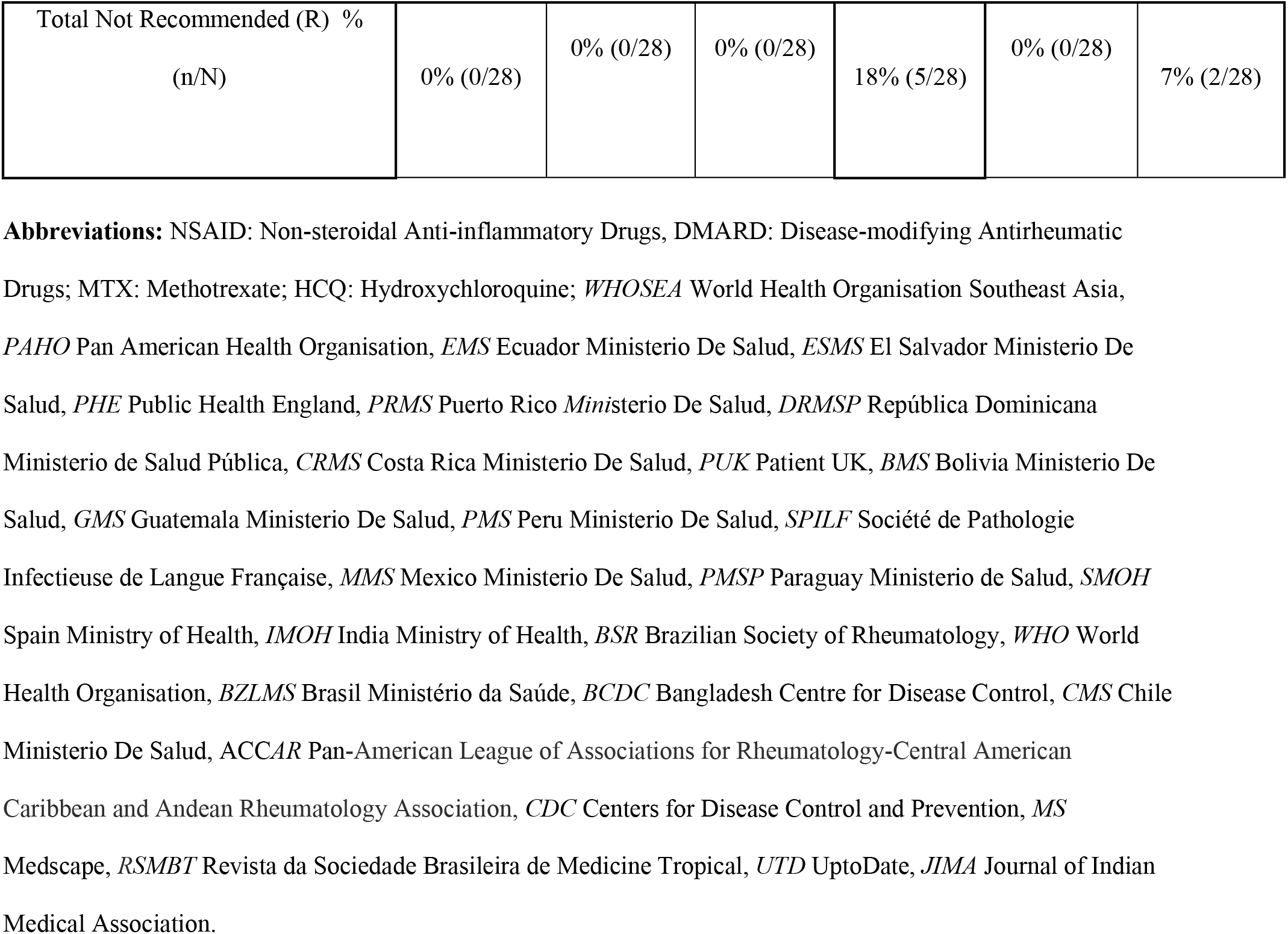
Summary of CMG recommendations for treatment of chronic sequalae. The table presents an overview of the main treatments recommended in the chronic phase in each guideline, and if a treatment was recommended to use, not recommended or if no advice was provided. R= recommended. RA= recommended to avoid. NS= not stated.

| Guidelines | Region | Year | Chronic interventions |  |  |  |  |  |
| --- | --- | --- | --- | --- | --- | --- | --- | --- |
|  |  |  | Analgesia |  |  | Steroids | DMARDs |  |
|  |  |  | Paracetamol | NSAIDs | Opioids |  | MTX | HCQ |
| ACCAR | Global | 2018 | NS | R | NS | NS | R | R |
| BCDC | Asia | 2017 | NS | NS | NS | NS | NS | R |
| BMS | Latin America | 2015 | NS | R | NS | R | R | NS |
| BSR | Latin America | 2017 | NS | R | R | R | R | R |
| BZLMS | Latin America | 2017 | R | R | R | R | NS | R |
| CDC | North America | 2018 | NS | R | NS | R | NS | NS |
| CMS | Latin America | 2018 | R | R | R | NS | R | NS |
| CRMS | Latin America | 2014 | R | R | NS | R | NS | NS |
| DRMSP | Latin America | 2014 | NS | NS | NS | RA | NS | NS |
| EMS | Latin America | 2014 | NS | R | NS | R | R | NS |
| ESMS | Latin America | 2014 | NS | R | NS | R | R | NS |
| GMS | Latin America | 2015 | NS | NS | NS | RA | NS | NS |
| IMOH | Asia | 2016 | NS | R | NS | NS | NS | R |
| JIMA | Asia | 2020 | NS | NS | NS | R | NS | R |
| MMS | Latin America | 2015 | R | R | NS | RA | NS | NS |
| MS | North America | 2019 | NS | NS | NS | RA | NS | NS |
| PAHO | Latin America | 2011 | NS | R | NS | R | R | RA |
| PHE | Europe | 2014 | NS | NS | NS | NS | NS | NS |
| PMS | Latin America | 2015 | R | NS | NS | NS | NS | NS |
| PMSP | Latin America | 2015 | NS | R | NS | R | R | NS |
| PRMS | Latin America | 2014 | NS | R | NS | NS | NS | NS |
| PUK | Europe | 2014 | R | NS | NS | NS | NS | NS |
| RSMBT | Latin America | 2020 | R | R | R | NS | R | R |
| SMOH | Europe | 2016 | R | NS | NS | R | R | NS |
| SPILF | Europe | 2015 | R | R | R | RA | R | NS |
| UTD | Global | 2020 | R | R | NS | R | R | RA |
| WHO | Global | 2017 | R | R | R | NS | R | NS |
| WHOSEA | Asia | 2008 | R | R | NS | R | NS | NS |
| Total Recommended (R) % (n/N) |  |  | 39%<br>(11/28) | 64%<br>(18/28) | 18%<br>(5/28) | 46%<br>(13/28) | 65%<br>(11/17) | 41%<br>(7/17) |

|  |  |  |  |  |  |  |
| --- | --- | --- | --- | --- | --- | --- |
| Total Not Recommended (R) %<br>(n/N) | 0% (0/28) | 0% (0/28) | 0% (0/28) | 18% (5/28) | 0% (0/28) | 7% (2/28) |

**Abbreviations:** NSAID: Non-steroidal Anti-inflammatory Drugs, DMARD: Disease-modifying Antirheumatic Drugs; MTX: Methotrexate; HCQ: Hydroxychloroquine; *WHOSEA* World Health Organisation Southeast Asia, *PAHO* Pan American Health Organisation, *EMS* Ecuador Ministerio De Salud, *ESMS* El Salvador Ministerio De Salud, *PHE* Public Health England, *PRMS* Puerto Rico Ministerio De Salud, *DRMSP* República Dominicana Ministerio de Salud Pública, *CRMS* Costa Rica Ministerio De Salud, *PUK* Patient UK, *BMS* Bolivia Ministerio De Salud, *GMS* Guatemala Ministerio De Salud, *PMS* Peru Ministerio De Salud, *SPILF* Société de Pathologie Infectieuse de Langue Française, *MMS* Mexico Ministerio De Salud, *PMSP* Paraguay Ministerio de Salud, *SMOH* Spain Ministry of Health, *IMOH* India Ministry of Health, *BSR* Brazilian Society of Rheumatology, *WHO* World Health Organisation, *BZLMS* Brasil Ministério da Saúde, *BCDC* Bangladesh Centre for Disease Control, *CMS* Chile Ministerio De Salud, *ACCAR* Pan-American League of Associations for Rheumatology-Central American Caribbean and Andean Rheumatology Association, *CDC* Centers for Disease Control and Prevention, *MS* Medscape, *RSMBT* Revista da Sociedade Brasileira de Medicina Tropical, *UTD* UptoDate, *JIMA* Journal of Indian Medical Association.

### Inclusivity

Most CMGs (89%, 25/28) ^9, 29, 31–35, 37–42, 44–53, 55^ mentioned vulnerable groups including children (75%, 21/28), ^9, 29, 31, 32, 37–43, 45, 46, 48–51, 53, 55^ pregnant women (68%, 19/28),^9, 29, 32, 33, 35, 38–45, 47–49, 51, 52, 53^ people aged >65 years (96% 27/28),^9, 29, 31–55^ people living with HIV (21%, 6/28)^29, 38, 40, 41, 49, 55^ and people living with comorbidities (61%, 17/28).^9, 29, 31, 32, 34, 36–38, 40, 41, 43, 44, 48–51, 54^ Many (61% (17/28)^30, 31, 33–37, 41, 42, 44, 45, 53–55^ provided some guidance for all of these groups; however, recommendations varied in their level of detail with only a minority giving specific supportive care guidance for pregnant women and children.

### Scope

All CMGs gave recommendations for the management of acute and chronic manifestations of chikungunya, although the level of detail varied (Table 2 and 3). There was considerable variation between CMGs in the recommendations for the management of patients with long-term sequelae and for different at-risk populations . Further, there were differences in the guidance addressing preventive measures of disease transmission both in the community and in hospital settings.

#### Acute Phase

Half of the CMGs (50%, 14/28)^9, 29, 37, 40, 41, 43, 45–47, 51–55^ stated that there was no efficacious antiviral treatment available for CHIKV and most (96%, 27/28) that management should be symptom-driven.^29–38, 40–55, 55^ Many (68%, 19/28) ^29, 32, 33, 35, 37, 38, 40–45, 47–53^ provided guidance on management of the acute phase, with administration at different health facility levels depending on disease severity: outpatient care (home based and at the primary care level), secondary level (district hospitals) and at the tertiary level (referral hospitals). The principles of outpatient management were generally consistent amongst the CMGs with recommendations including rest (39%, 11/28), ^32, 33, 36, 38, 40, 46–48, 52, 53, 55^ hydration (43%, 12/28), ^29, 31–33, 36–38, 41, 46, 52, 53, 55^ cold compresses (11%, 3/28), ^38, 45, 51^ antihistamines (39%, 11/28)^32, 38, 40, 42, 44, 45, 49, 53–55^ and analgesia (96% 27/28). A minority (25%, 4/12)^29, 37, 38, 52, 53^ advised the need to assess the patient’s hydration status to assess whether intravenous fluid was required, one (8%, 1/12)^53^ advised suspending diuretics.

Fifty-four percent of CMGs (15/28)^33, 35, 37, 38, 40–45, 47–49, 51, 52^ recommended hospitalisation for severe cases; however, only a minority (39%, 11/28)^9, 29, 33, 34, 37, 38, 41, 46, 51, 52, 54^ gave guidance regarding the clinical management of severe cases. The CMGs (54%, 15/28)^33, 35, 37, 38, 40–45, 47–49, 51, 52^ providing guidance regarding hospitalisation criteria gave several indications including: any signs of haemodynamic instability (46%, 13/28), ^33, 35, 38, 41–45, 47–49, 51, 52^ or atypical chikungunya (36%, 10/28),^29, 33, 35, 38, 41, 43, 47–49, 52^ severe pain unresponsive to analgesia (25%, 7/28),^35, 37, 38, 41, 42, 44, 45^ signs of haemorrhage (46%, 13/28)^29, 33, 35, 38, 41–45, 47–49^ and signs of decompensation from underlying comorbidities (25%, 7/28).^29, 33, 35, 38, 43, 47, 49^ Supportive care recommendations included use of intravenous fluids to treat shock (55%, 6/11),^29, 37, 45, 47, 48, 52^ haemodynamic monitoring (55%, 6/11),^29, 33, 34, 45–47^ blood components (18%, 2/11), ^37, 45^ intensive care support as required (9%, 1/11)^9^ and immunoglobulins in chikungunya-related polyneuropathy (4%, 1/28).

#### Antimalarials

None of the CMGs advocated for use of empiric antimalarials, despite some CMGs (10%, 2/21)^9, 44^ advising malaria should be considered in the differential diagnosis. However, CMGs did discuss the use of antimalarial chloroquine derivatives for the treatment of chronic manifestations (24%, 4/17).^33, 37, 44, 45^

#### Analgesia

All CMGs recommended some form of analgesia with 75% (21/28)^29, 31–33, 35, 37, 40–48, 52–55, 55^ recommending paracetamol as first line treatment for pain and its antipyretic properties. Some (36%, 10/28)^9, 33, 35, 40, 47, 48, 52– 55^ advised escalating to opiates, tramadol, or codeine alone or in combination with paracetamol, for uncontrolled pain. Two (7%, 2/28)^33, 51^ recommended dipyrone for mild pain. There was wide and contradictory advice given regarding non-steroidal anti-inflammatory drugs (NSAIDs) in the acute phase. Half (54%, 15/28) advised considering the use of NSAIDs.^29, 31, 32, 34, 36–42, 44, 47, 48, 55^ However, 75% (21/28) advised avoiding salicylates in adults during the acute phase due to risk of haemorrhage,^9, 29, 32–35, 37–44, 47–53^ and 40% (11/28) advised against NSAIDs,^9, 33, 35, 43, 45, 46, 49, 51–54^ while one CMG stated that NSAIDs should not be avoided, citing a lack of evidence.^32^ Some (29%, 8/28)^9, 29, 35, 41, 46, 48–50^ recommended excluding co-infection with dengue prior to NSAID administration.

#### Corticosteroids

The recommendations for corticosteroids were also heterogenous. A third (36%, 10/28)^32, 35, 41, 44, 46–48, 50, 53, 54^ advised a short course of corticosteroids if not responding to analgesia. A number of indications were given for corticosteroid, including severe joint pain refractory to NSAIDs or other analgesia (80%, 8/10);^32, 35, 41, 44, 46–48, 53^ highly inflammatory forms (30%, 3/10); ^32, 53, 54^ disabling arthritis (40%, 4/10)^32, 41, 44, 54^ or when contraindications against NSAIDs (10%, 1/10).^53^ Prednisolone was most recommended (50%, 5/10), ^32, 41, 46, 53, 54^ with CMGs advising adult dosage ranging from 10mg to 20mg per day (60%, 3/5) based on clinical judgment,^32, 46, 53^ escalating to 0.5mg/kg/day (80%, 4/5)^32, 41, 53, 54^ for severe cases. Four CMGs (80%, 4/5)^32, 41, 53, 54^ gave corticosteroid duration advise, ranging, from 5 days (60%, 3/5)^32, 41, 53^ with a weaning period of 10 days to 1-2 months for severe cases (40%, 2/5)^32, 54^ with a correspondingly longer weaning period. In contrast, two CMGs (40%, 2/5) stated that the duration of corticosteroid use should not exceed one month,^32, 53^ with one citing the SPILF guideline.^9^ Although the majority (80%, 4/5)^32, 41, 53, 54^ advised on the need for tapering steroid doses, only one CMG gave a justification, stating that there was a risk of rebound symptoms if abruptly withdrawn.^54^ The remaining CMGs (50%, 5/10) did not specify corticosteroid dose or type.^35, 44, 47, 48, 50^ In contrast to these recommendations, 43% (12/28)^9, 29, 34, 37, 38, 40, 43, 45, 49, 51, 52, 55^ advised against the use of steroids in the acute phase. Only a minority gave justifications for avoidance, stating either a lack of evidence (8%, 1/12),^34^ lack of short-or long-term benefit regardless of form of administration (8%, 1/12)^52^ or a risk of rebound symptoms (8%, 1/12).^55^ Furthermore, one CMG [cite] had contradictory recommendations within its guideline, advising the use of short-term corticosteroids for individuals with refractory pain in the acute phase, while also advising against the use of systemic corticosteroids in the acute phase.^29^

A minority of CMGs (14%, 4/28)^29, 32, 33, 54^ also indicated the use of steroids in the subacute phase to treat symptoms refractory to NSAIDs, moderate pain and for arthritis/tenosynovitis. Three of these CMGs (75%, 3/4)^32, 33, 54^ advised that prednisolone was first-line, and that steroids use should be limited to one month. One CMG (25%, 1/4)^33^ provided recommendations on how to assess improvement (ability to walk without assistance; satisfactory pain control) to guide dose and duration.

#### Chronic Phase

Most CMGs (93%, 26/28)^9, 29, 32–55^ addressed the management of long-term sequelae, based on the principles of managing more common inflammatory arthropathies such as rheumatoid arthritis (RA). The recommendations included analgesia, corticosteroids, disease modifying anti-rheumatic drugs (DMARDs) and antimalarial chloroquine derivatives. Some CMGs (18%, 5/28)^9, 33, 42, 51, 54^ advised using quantitative scoring measures (visual scales, clinical scores and structured questionnaires) to measure outcomes such as pain, joint involvement, quality of life and functional capacity in adults. The most common scale recommended to assess severity and to monitor the efficacy of treatment was a visual analogue scale (VAS) (80%, 4/5).^9, 33, 51, 54^ Other scales recommended were Routine Assessment of Patient Index Data 3 (RAPID3), Disease Activity Score-28 (DAS28) and Douleur Neuropathique 4 (DN4) to assess the functional impact of pain and level of neuropathic pain, respectively.^9, 54^

##### Analgesia

Most CMGs (86%, 24/28)^9, 29–33, 35, 36, 39–44, 46–54^ advised managing chronic pain using analgesia, primarily NSAIDs (75%, 18/24), ^9, 29, 32, 33, 35, 41–44, 46–50, 50–52, 54^ paracetamol (45%, 11/24)^9, 31–33, 35, 39, 40, 42, 49, 52, 54^ and opiates (21%, 5/24).^9, 33, 51, 52, 54^ A minority (13%, 3/24)^51, 53, 54^ gave guidance regarding the duration of treatment which ranged from reassessing after four, ^51^ eight^54^ to several weeks.^53^ The rest (92%, 22/24) lacked clarity regarding the length of time analgesia should be continued.^9, 29, 35, 36, 38, 39, 46–49, 49, 50, 52^

#### Corticosteroids

Almost half the CMGs (46%, 13/28) advised giving steroids in the chronic phase.^29, 32, 36, 37, 39, 41–44, 47, 51, 52^ The most common indication given for administration of corticosteroids was disabling peripheral arthritis refractory to other treatments (62%, 8/13),^29, 33, 36, 39, 41, 44, 47, 52^ followed by neuropathic symptoms (8%, 1/13).^51^ Moreover, to those who experience arthralgia, arthritis, tendinitis, or bursitis with evidence of severe synovitis, joint swelling and persistent elevation of inflammatory markers (8%, 1/13).^32^ Most recommended prednisolone (31%, 4/13),^32, 33, 41, 51^ most (75%, 3/4) specifying a dosage of 0.5mg/kg/day.^32, 33, 41^ There was considerable variation in the recommendations regarding duration, with CMGs advising courses of five,^32^ ten,^41^ 21,^33^ or 28 days.^37^ One CMG (8%) advised using 5 to 20 mg/day prednisone for musculoskeletal and neuropathic symptoms for 6 to 8 weeks with a weaning period to avoid symptom recurrence.^51^ For administration, although most CMGs advised oral steroids (31%, 4/13),^33, 37, 39, 51^ a few advised that local intra-articular injections could be used (15%, 2/13).^39, 47^ A minority of CMGs (18%, 5/28)^9, 34, 38, 49, 55^ advised against the use of corticosteroids in the chronic phase giving reasons such as the risk of rebound symptoms (20%, 1/5)^55^ and lack of published evidence (20%, 1/5).^34^ The rest of the CMGs did not give a justification for avoidance (60%, 3/5).^9, 38, 49^

#### DMARDs

Over half of the CMGs (61%, 17/28)^9, 29, 32, 33, 37, 39, 41, 43–48, 51–54^ gave guidance on the use of DMARDs in the chronic phase; yet there was variation on which DMARD was first line. Methotrexate was recommended as first line therapy by most (65%, 11/17);^9, 29, 32, 39, 41, 43, 46–48, 52, 53^ whereas others (24%, 4/17) recommended chloroquine/ hydroxychloroquine.^33, 44, 45^ One (6%, 1/17) recommended that methotrexate should be used for inflammatory joint disease (moderate or severe disease affecting >5 joints) and hydroxychloroquine reserved for less severe forms.^54^ Another (6%, 1/17) noted that there was a lack of data comparing the efficacy of methotrexate and hydroxychloroquine, but recommended hydroxychloroquine as it the safer choice for its anti-inflammatory and possible antiviral effects.^33^ Two CMGs (12%, 2/17) recommended methotrexate either in combination with another DMARD, such as sulfasalazine or chloroquine.^46, 51^ One CMG (6%, 1/17) divided chronic manifestations into post-chikungunya rheumatoid arthritis (methotrexate first line), post-chikungunya spondyloarthritis (NSAIDS first line) and post-chikungunya undifferentiated polyarthritis (NSAIDs first line; corticosteroids second line).^9^ Five CMGs (18%) provided guidance for neuropathic pain management of using amitriptyline, pregabalin, gabapentin and carbamazepine.^9, 33, 45, 51, 54^

#### Vulnerable populations

##### Pregnant women

Most (75%, 21/28) addressed management during pregnancy.^9, 29, 31–33, 35, 36, 38–45, 47–49, 51–53^ Yet, limited CMGs (29%, 6/21) ^9, 38, 41, 43, 52, 53^ gave guidance on CHIKV symptom control during pregnancy, advising to use paracetamol (67%, 4/6).^9, 38, 52, 53^ Moreover, to consider amoxicillin in febrile (>38.5C) women (17%, 1/6) ^9^ and to avoid NSAIDs and aspirin (50%, 3/6) ^9, 52, 53^ due to the risks of closure of the ductus arteriosus, fetal renal failure and risk of intrauterine death [cite]. Some CMGs (57%, 12/21) recommended referral to health services for monitoring of mother and child, but the advice varied.^9, 29, 35, 38, 40, 47–49, 51, 52^ One (8%, 1/12)^35^ recommended admitting all pregnant women with suspected chikungunya if in the last trimester; whereas one (8%, 1/12) specified from 38 weeks.^48^ In contrast, two CMGs (16%, 2/12) ^33, 49^ recommended daily monitoring of pregnant women with suspected chikungunya; three (25%, 3/12) recommended obstetric referral if in the final trimester.^9, 52, 53^ Delaying delivery beyond the highly viraemic stage to try to prevent mother-to-child transmission (MTCT) was advised in four (33%, 4/12). One (8%, 1/12) ^9^ advising use of tocolytics, one (8%, 1/12)^35^ postponement of elective caesarean section. Further, four CMGs (19%, 4/21) advised that caesarean sections did not prevent mother-to-child transmission (MTCT).^9, 38, 45, 49^

##### Children

Most CMGs (79%, 22/28) identified children and neonates as having a higher risk of developing severe CHIKV infection and advised hospital referral, but their criteria varied.^9, 29, 31, 33, 35, 37–40, 40–55^ Four (18%, 4/22) ^9, 41, 52, 53^ advised inpatient monitoring for signs of infection of neonates born to mothers with suspected chikungunya for seven days. The guidance changed for neonates born to mothers with confirmed infection, with three CMGs (14%, 3/22) advising that inpatient monitoring should be five days, ^9, 52, 53^ one at least seven days.^41^ One (5%, 1/22) recommended that symptomatic neonates should be managed in the neonatal intensive care unit.^38^ Four (18%, 4/22) addressed breastfeeding, stating there was no risk of transmission through breastmilk.^38, 44, 45, 47^ Four CMGs (18%, 4/22) advised infants at risk of CHIKV infection younger than 12 months to be admitted to hospital for observation.^29, 37, 40, 45^ Four (18%, 4/22) advised children under two years old to be followed up daily in a primary care facility during the acute phase [cite]. Nine CMGs (32%) specified the risk of Reye’s syndrome associated with aspirin use in children younger than 12 years old **[cite].** Four (18%, 4/22)^9, 29, 52, 53^ advised against NSAID administration in children under the age of 3 months, and three (11%) against codeine use in children younger than 12 years.^9, 52, 53^ One CMG (4%) advised against use of dipyrone in infants younger than three months or weighing less than 5kg.^54^

##### Older adults and those with comorbidities

Although 96% (27/28) of CMGs^9, 29, 30, 32, 34–51, 53–56^ included advice for older adults (CMGs generally defined as those aged over 60 or 65 years) and those with comorbidities, this advice was limited in scope. Two (22%, 2/9) stated that people over 60 years old had a 50-times higher mortality risk compared to younger adults.^41, 43^ While 81% (22/27)^9, 29, 31, 32, 35–39, 41, 42, 44, 46–53, 55^ advised that older adults were at increased risk of severe/atypical disease and death, only seven (26%, 7/27)^37, 41, 42, 45, 48, 53^ recommended referral to hospital for monitoring. One CMG (11%, 1/9) stated that in people over 65 years old CHIKV infection could cause complications and lead to dementia, paralysis and kidney disease.^44^ Over half of the CMGs (61%, 17/28)^9, 29, 31, 32, 34, 36, 38, 40, 43, 44, 48–52, 54^ advised that people with chronic conditions such as diabetes, hypertension or heart disease were at higher risk of developing severe/ atypical disease or deterioration due to decompensation of their pre-existing condition. Of these, nine (81%, 9/11) suggested having a lower threshold for hospital referral, and three close monitoring of these patients, and adult over 65 years old. In keeping with general guidance, five CMGs (45%, 5/11) advised prescribing NSAIDs with caution in patients with comorbidities due to risk of renal impairment and bleeding risk.^33, 38, 40, 48, 54^

#### Prevention of onward transmission

Most CMGs (71%, 20/28)^9, 29, 31, 32, 35, 37, 38, 40–51, 53^ gave advice regarding prevention of transmission. Recommendations included the use of mosquito repellents (50%, 10/20),^9, 31, 35, 40, 41, 43, 44, 47, 50, 51^ protective clothing (35%, 7/20), ^31, 40, 41, 43, 50, 51^ mosquito nets (60%, 12/20), ^9, 37, 40–44, 47, 49–51, 53^ and isolation (25%, 5/20) of the patient and those in proximity to the patient. It was recommended to continue these measures throughout the febrile phase of illness to reduce risk of transmission. In contrast, two CMGs stated that there was no requirement to segregate the infected patient in a household.^44, 45^ Only three CMGs (15%, 3/20) advised on the risk of blood-borne transmission^9, 38, 46^ with one specifying highest risk within the first five days of symptomatic infection.^38^ Two (10%, 2/20) highlighted risk of transmission via organ/tissue transplantation.^38, 46^ Seven CMGs (35%)^9, 29, 44, 45, 48, 50, 51^ recommended vector control measures around the hospital/homes of infected patients, using insecticides,^9, 48^ fumigation^29^ and eradication of breeding sites.^9, 50^ Some (65%, 13/20) advised informing public health authorities. ^29, 31, 35, 40–42, 44, 45, 47^

## Discussion

This review highlights the limited availability of high-quality CMGs for chikungunya globally. In the CMGs identified there were heterogenous recommendations on supportive care and treatments. Although there was a consensus in the guidelines on the symptomatic treatment for acute non-severe illness, there was a general lack of detailed supportive care guidance regarding patients with acute severe disease.

Furthermore, there were significant differences in the guidance around corticosteroids, with certain CMGs advocating for their use in the acute phase, while a third advised that the acute phase was a contraindication. The duration of steroid treatment for both acute and chronic disease was another point of contention between the CMGs. The evidence base around corticosteroid use in acute illness is uncertain with studies limited in size and a scarcity of RCTs investigating corticosteroid use in acute CHIKV.

The evidence base around corticosteroid use in acute CHIKV is uncertain. One prospective randomized parallel group study of 120 patients with acute CHIKV in South India demonstrated that the addition of corticosteroid to NSAIDs reduced pain and improved quality of life and advocated for combination treatment in acute illness.^57^ Another small study of 19 cases observed an improvement in mobility with short term corticosteroids in acute CHIKV, however noted that there was a risk of rebound symptoms after treatment cessation.^58^ Several reviews advised against the use of corticosteroids citing risks such as rebound symptoms and immunosuppression causing potential disease exacerbation.^59, 60^ It can be reasonably assumed that the clinical guidelines are contradictory and lack clarity due to a scarcity of research into the use of corticosteroids in acute chikungunya infection.

The joint pain caused by CHIKV infection may be debilitating, which can limit even the simplest daily activities. Polyarthralgia is recurrent in 30 to 40% of infected individuals and may persist for years.^61^

Furthermore, CHIKV infection can lead to death either through the infection and its associated complications or by triggering a decompensation in patients with pre-existing co-morbidities.^62^ The risk of prolonged sequelae in populations in lower resourced settings especially can have a profound impact on livelihoods, with a wider socio-economic impact on individuals, their families and the wider society. Considering the high number of people affected and at risk of CHIKV infection, the scarcity and heterogeneity as well as the sometimes contradictory treatment recommendations in the CMGs available for chikungunya are reasons for concern.

Most CMGs included guidance on the treatment for pregnant women and children and there was general consensus that this group were at higher risk of severe infection. Yet, guidance regarding symptomatic treatment was limited and there were variations in the guidance around referral criteria to health services. Moreover, there were wide variations in risk and need for monitoring of infection in young children, and in the treatment recommendations and duration. Furthermore, only a minority of CMGs mentioned potential factors to mitigate the risks of MTCT during delivery. For children, although many CMGs identified that they were a high-risk group, only a minority gave guidance on hospitalisation criteria and advised on symptom control. The CMGs were also limited in specific advice for older people and for those with co-morbidities, both of whom are at higher risk of more severe disease. Given the substantial risk CHIKV infection presents to neonates, the lack of clear guidance around reducing MTCT is a concern. Although there are novel approaches to prevent the risk of MTCT, such as anti-CHIKV hyperimmunoglobulins, there is currently no approved treatment ^63–65^ and published studies are scarce.

Furthermore, there was considerable variation in the guidance for treating and managing long-term chronic, often debilitating, sequelae. The evidence into effective treatments is limited, and symptomatic treatment without appropriate individual follow-up, whilst crucial to the quality of life of many patients, has not been found to have an effect in diminishing mortality.^66^ A systematic review of five RCTs with small sample sizes found that the evidence on treatments was insufficient from a safety or efficacy point of view.^67^

Although most CMGs provided recommendations for post-acute follow-up care and treatment of chronic complications, the recommendations were heterogenous and with limited evidence provided to support them. There was variation in the recommendations of DMARDs for the management of chronic chikungunya, particularly between hydroxychloroquine and methotrexate. One study examined combination DMARD therapy versus hydroxychloroquine treatment in 72 patients with post-chikungunya arthritis and found that a combination of DMARDs were superior to hydroxychloroquine monotherapy treatment with improvements in disability, reduction in pain and disease activity.^68^ Despite acknowledging this lack of benefit, however four CMGs recommended hydroxychloroquine as a first line DMARD.^33, 37, 44, 45^ Existing interventional clinical research studies are limited in size as highlighted in Martí-Carvajal et al., with a lack of standardised methodologies, the ability to conduct meta-analyses is restricted, thus limiting our evidence base in determining the most effective therapies for treating chronic manifestations of CHIKV infection.^67^ The paucity of clear guidance is a disservice to patients, particularly given that most patients developing acute severe illness and complications fall within vulnerable groups. Our data highlights a need for robust RCTs with adequate statistical power to identify best supportive care and new treatments to improve short and long term CHIKV outcomes.

This review is not without limitations. Despite a systematic search, additional local guidelines not retrieved by our searches may exist. Approximately half of the included CMGs were in a language other than English, and although these were assessed by a reviewer with good knowledge of that language, there may have been slight nuances lost in translation. Furthermore, the AGREE-II tool ^26^ assess methodological aspects relevant to guideline development, but not the validity of the clinical management recommendations themselves. Whilst many of the CMGs scored poorly in the rigour of development domain, conclusions about the validity of the clinical guidance made can therefore not be derived from these scores.^26^ Despite these limitations, this review identifies concerning gaps and disparities within the CMGs. Firstly, there is an issue of accessibility with the two highest quality CMGs identified in this review not being freely available. One is only available via paid subscription^69^ and the other a national guideline in Spanish.^49^ Developing CMGs is resource intensive, and for infectious diseases with rapidly changing epidemiology, requires systems for regular reviews of the literature, updating and re-dissemination. CHIKV disproportionally impacts on lower resourced settings, where such resources may not be readily available. Further, other infections may take priority when there is international pressure and/or funding to develop research and guidelines (e.g., SARS-CoV-2, HIV, malaria). International high-quality CMGs can fill this gap, if they are readily accessible, and can easily by adapted and adopted by local settings during outbreaks. This may also help ensure there are resources available to incorporate new evidence and disseminated this via international platforms. The WHO new living Covid-19 review is an example of this.

## Conclusion

There is a lack of high-quality CMGs detailing supportive care guidance in chikungunya globally, particularly for those at risk of severe illness. Given the risks that CHIKV infection pose globally and in particular to vulnerable groups such as children, individuals older than 65 years and those with co- morbidities, it is essential that existing guidelines are updated and adapted to include recommendations for people with severe illness. Chikungunya is an illness predominantly affecting populations in lower-resourced settings, with profound impact on quality of life and livelihoods. Further research is needed into effective treatments and vaccines, to generate evidence to inform high quality CMGs and improve patient and epidemic outcomes. Investment in a ‘living review’ framework, for international CMGs that can be readily adopted by local settings is recommended to improve access to inclusive, up to date, evidence-based treatment guidelines.

## Data Availability

All relevant data are within the manuscript and its Supporting Information files.

## Author’s contributions

AD, VC, LS, SL, EH, STJ, EW, MM, IR developed the study protocol. EH, AD carried out the database search with input from MM, EW and IR. EW, MM, IR, AD, EC conducted the grey literature search. EW, RJ, AD, MM screened articles for inclusion. AD, EW, IR, RJ, MM extracted the data and completed the risk of bias analysis. EW, MM, RJ, SL, and IR led on data analysis and presentation of the results. All co-authors informed the interpretation of the findings. EW led on writing the manuscript with inputs from LS, IR, MM, DD, RN, PPJ, ES and KG. LS, PH, TF, LB and STJ provided overall supervision, leadership and advice. PH, HG, STJ, TF, PWH, LS, AD, and LB conceptualised the project. All authors reviewed and approved the final version of the manuscript.

## Acknowledgements

Thanks to the ISARIC Global Support Centre for logistical and administrative support, especially to Sarah Moore and Romans Matulevics for invaluable support for this project.

## Ethical approval

Not required

## Data sharing

All of the data is presented in the manuscript and supplementary material.

## Transparency statement

The lead author (the manuscript’s guarantor) affirms that the manuscript is an honest, accurate, and transparent account of the study being reported; that no important aspects of the study have been omitted; and that any discrepancies from the study as originally planned and registered have been explained.

## List of abbreviations

CHIKV: Chikungunya Virus
CMG: Clinical Management Guideline
DMARD: Disease Modifying Anti-Rheumatic Drug
PAHO: Pan American Health Organisation
WHO: World Health Organisation
AGREE: Appraisal of Guidelines for Research and Evaluation II
PROSPERO: The International Prospective Register of Systematic Reviews
SARS-CoV-2: severe acute respiratory syndrome coronavirus 2
COVID-19: Coronavirus Disease
PRISMA: Preferred Reporting Items for Systematic Reviews and Meta-Analyses
CADTH: Canadian Agency for Drugs and Technologies in Health
ISARIC: International Severe Acute Respiratory and Emerging Infection Consortium
WHO: World Health Organisation
PAHO: Pan American Health Organisation
HIV: Human Immunodeficiency Virus
IQR: Interquartile range
WHOSEA: World Health Organisation Southeast Asia
ACCAR-Pan: American League of Associations for Rheumatology-Central American Caribbean and Andean Rheumatology Association
BCDC: Bangladesh Centre for Disease Control
BMS: Bolivia Ministerio De Salud
BSR: Brazilian Society of Rheumatology
BZLMS: Brasil Ministério da Saúde
CDC: Centers for Disease Control and Prevention
CMS: Chile Ministerio De Salud
CRMS: Costa Rica Ministerio De Salud
DRMSP: República Dominicana Ministerio de Salud Pública
EMS: Ecuador Ministerio De Salud
ESMS: El Salvador Ministerio De Salud
GMS: Guatemala Ministerio De Salud
IMOH: India Ministry of Health
JIMA: Journal of Indian Medical Association
MMS: Mexico Ministerio De Salud
MS: Medscape
PHE: Public Health England
PMS: Peru Ministerio De Salud
PMSP: Paraguay Ministerio de Salud
PRMS: Puerto Rico *Mini*sterio De Salud
PUK: Patient UK
RSMBT: Revista da Sociedade Brasileira de Medicine Tropical
SMOH: Spain Ministry of Health
SPLIF: Société de Pathologie Infectieuse de Langue Française
UTD: UptoDate
NSAIDs: Non-Steroidal Anti-inflammatory Drugs
RAPID3: Routine Assessment of Patient Index Data 3
DAS28: Disease Activity Score-28
DN4: Douleur Neuropathique 4
DMARD: Disease Modifying Anti-rheumatic Drugs
MTCT: mother-to-child transmission
RCT: Randomised Control Trial
MTX: Methotrexate
HCQ: Hydroxychloroquine R-Recommended
NS: Not Stated
RA: Recommended to avoid

